# Intrathecal Nicardipine for Cerebral Vasospasm Post Subarachnoid Hemorrhage–a Retrospective Propensity-Based Analysis

**DOI:** 10.1101/2020.08.31.20185181

**Authors:** Ofer Sadan, Hannah Waddel, Reneé Moore, Chen Feng, Yajun Mei, David Pearce, Jacqueline Kraft, Cederic Pimentel, Subin Mathew, Feras Akbik, Pouya Ameli, Alexis Taylor, Lisa Danyluk, Kathleen S. Martin, Krista Garner, Jennifer Kolenda, Amit Pujari, William Asbury, Blessing Jaja, R. Loch Macdonald, C. Michael Cawley, Daniel L. Barrow, Owen Samuels

**Affiliations:** Department of Neurology and Neurosurgery, Division of Neurocritical care, Emory University School of Medicine, Atlanta, Georgia, USA; Department of Biostatistics and Bioinformatics, Biostatistics Collaboration Core, Emory University, Atlanta, Georgia, USA; H. Milton Stewart School of Industrial and Systems Engineering, Georgia Institute of Technology, Atlanta, Georgia, USA; Neuroscience ICU, Emory healthcare, Atlanta, Georgia, USA; Emory University School of Medicine, Atlanta, Georgia, USA; Department of Clinical Pharmacy, Emory Healthcare, Atlanta, Georgia, USA; Department of Genetics and Development, Toronto Western Hospital, University Health Network, Toronto, ON, Canada; Department of Neurological Surgery, University of California San Francisco, Fresno Campus, Fresno, California, USA; Department of Neurosurgery, Emory University Hospital and School of Medicine, Atlanta, Georgia, USA

**Keywords:** Subarachnoid hemorrhage, delayed cerebral ischemia, vasospasm, calclium channel blocker, intrathecal administration

## Abstract

**Objectives:** Cerebral vasospasm and delayed cerebral ischemia (DCI) contribute to poor outcome following subarachnoid hemorrhage (SAH). With the paucity of effective treatments, we describe our experience with intrathecal (IT) nicardipine for this indication.

**Methods:** Patients admitted to Emory University Hospital Neuroscience ICU between 2012-2017 with non-traumatic SAH, either aneurysmal or idiopathic, were included in the analysis. This patient cohort was compared using a propensity-score model to patients in the SAH international trialist (SAHIT) repository who did not receive intrathecal nicardipine. The primary outcome was DCI. Secondary outcomes were long-term functional outcome and adverse events.

**Results:** The analysis included 1,351 patients, 422 of whom were diagnosed with cerebral vasospasm and treated with IT nicardipine. When compared with patients with no vasospasm (n=859) the treated group was younger (51.1±12.4 vs. 56.7±14.1, p<0.01), had a higher World Federation of Neurological Surgeons score (WFNS), modified Fisher grade, and more likely to undergo clipping of the ruptured aneurysm as compared to endovascular treatment (30.3% vs. 11.3%, p<0.01). Treatment with IT nicardipine decreased daily mean transcranial Doppler velocities in 77.3% of the treated patients. When compared to patients not receiving IT nicardipine, treatment was not associated with an increase rate of bacterial ventriculitis (3.1% compared with 2.7%, p>0.1) yet higher rates of ventriculoperitoneal shunting were noted (19.9% vs. 8.8%, p<0.01). In a propensity score comparison to the SAHIT database, the odds ratio to develop DCI with IT nicardipine treatment was 0.61 with 95% CI[0.44-0.84], and to have a favorable functional outcome (mRS≤2) was 2.17[1.61-2.91].

**Conclusions:** IT nicardipine was associated with improved outcome and reduced DCI compared with propensity matched controls. There was an increased need for permanent CSF diversion but no other safety issues. This data should be considered when selecting medications and treatments to study in future randomized controlled clinical trial for SAH.

## Introduction

Non-traumatic subarachnoid hemorrhage (SAH) is an uncommon, but often devastating, cause of stroke that disproportionately causes high mortality and morbidity. In-hospital mortality rate remains very high, approximately 12-20%, despite advancements in care during the last few decades.^1–3^ In survivors, functional outcomes are limited, resulting in 40% of the patients unable to return to their prior line of work.^4^ One of the common and often devastating neurologic effects of SAH patients is angiographic (cerebral) vasospasm and delayed cerebral ischemia (DCI).^5^ Diagnostic imaging can detect cerebral vasospasm in up to 70% of SAH patients using CT angiogram (CTA) or digital subtraction angiography (DSA). Symptomatic vasospasm, however, is reported to develop in 20-30% of patients with SAH,^5,6^ and a similar rate is cited for radiographic ischemic changes consistent with DCI.^7^ Cerebral vasospasm and DCI both correlate to functional and cognitive outcome of patients with SAH,^8–10^ as well as with systemic complications during the ICU stay.^11^

Angiographic vasospasm and the development of DCI have a complex and incompletely understood inter-relationship. Conventional wisdom suggests that vasospasm of the major proximal intracranial vasculature (macrovascular vasospasm) of the anterior and posterior circulation leads to DCI by reducing cerebral perfusion. Evidence suggests, however, that intracranial vasospasm also occurs in the micro-vasculature, and in the absence of large arterial (macro) vasospasm, which may be alleviated by dihydropyridine drugs.^12^ Indeed, nimodipine, the only prophylactic treatment with Level I evidence to support its use, results in improved patient outcomes, with minimal to no effect on angiographic vasospasm.^13^

Nicardipine, another dihydropyridine calcium antagonist, with similar pharmacologic properties to nimodipine, did not improve clinical outcome in a phase III trial when administered intravenously to patients with SAH in randomized controlled trials. Prophylactic intravenous nicardipine, however, did reduce angiographic vasospasm without demonstrating clinical benefit in reducing DCI.^14,15^ One theory to explain the reduction in angiographic vasospasm without improved clinical outcome was that systemically administered nicardipine caused hypotension that was detrimental. One strategy to avoid systemic side effects is intrathecal delivery via an external ventricular drain (EVD), although there is conflicting data as to the efficacy of this approach.^16–23^

In contrast to a purely preventive approach for all-comers, an alternative strategy is early recognition and intervention. This way, treating cerebral vasospasm occurs only when it is diagnosed. Hence, patients who do not develop angiographic vasospasm will not be exposed to the treatment and its potential side effects.

In this report, we summarize the efficacy and safety data of patients with SAH treated therapeutically for cerebral vasospasm. We compared the findings to a known established international registry of SAH trials and single center experiences, namely the SubArarchnoid Hemorrhage International Trialists (SAHIT) database.^24^ A propensity score-based weighted analysis was utilized. We hypothesized that when compared with a similar SAH patient population, the use of intrathecal (IT) nicardipine results in lower rates of radiographic DCI and improved long-term functional outcome.

## Methods

A retrospective clinical data collection of hospital records from patients with SAH admitted to our institution and discharged between January 1, 2012, and December 31, 2017 form the basis of this cohort. The institutional review board at Emory University, Atlanta, GA, approved the data collection and quality assurance analysis, and waived the need for patient consent. All patients with a discharge diagnosis of SAH during this time frame were screened. Exclusion criteria included a definite non-aneurysmal etiology (e.g. traumatic SAH, arterio-venous malformation etc.), while aneurysmal and idiopathic (angio-negative) SAH were included.

Detailed demographic, clinical assessments, and clinical data, including transcranial Doppler (TCD), and imaging were obtained from the individual chart review. TCDs were measured daily, 7 days a week. If TCD was not available (e.g. lack of bone sonographic windows, machine availability), surveillance CTA was used at a frequency determined by the clinical team. Clinically relevant cerebral vasospasm was defined as radiographic evidence (CTA and/or TCDs and/or DSA) that the clinical team (neurocritical care, vascular neurosurgery) decided justified an intervention (such as induced hypertension, cerebral perfusion pressure management, IT nicardipine etc.). There was substantial variability in the diagnosis of vasospasm, and we did not identify specific numerical thresholds in this retrospective analysis. We defined a subgroup of patients who developed vasospasm (+vasospasm, compared with –vasospasm, hence no clinically relevant vasospasm determination in real time), as well as a third sub-group of patients who were treated with IT nicardipine (all within the positive vasospasm group).

Brain and vascular imaging were assessed by two study neurologists who were blinded to avoid bias. Raters were not aware of the vasospasm intervention, severity of vasospasm nor clinical outcome. Radiological DCI was defined as any new ischemic lesion that was not apparent on a scan 24-48 hours after aneurysm repair procedure, and cannot be attributed to another etiology, within six weeks of the bleeding event.^7^ In the case of any disagreement among the evaluators, the findings were discussed to reach a consensus.

The TCD data was available for analysis from November 2014 onwards (when the TCD reports became digitally stored). TCDs were acquired daily by a single operator (n=356). The effect of IT nicardipine was measured as the daily change in mean velocities and Lindegaard ratio (LR).^25^ Response index was defined as the ration between the average TCD velocity in the MCA territory 24h post treatment and the velocity before treatment was initiated. The supplementary information reports the CTA information.

CSF was obtained routinely 3 times a week for surveillance chemistry and cultures. Bacterial ventriculitis was defined as the presence of a positive CSF culture that warranted a complete course of antibiotic treatment. Contamination, specifically an immediate sterile repeated culture, resulted in holding antibiotic care, was not defined as ventriculitis.

Outcome measures and complications were derived from the chart review. Long-term functional outcome defined by modified Rankin scale (mRS) was obtained from the clinic visit following the acute admission.

A comparative cohort was generated from the SAHIT database. Anonymized patient-level information was aggregated by selecting the specific datasets from the SAHIT database that met the following criteria: patients treated after International Subarachnoid Aneurysm Trial (ISAT)^26^, to avoid the influence of bias caused by increased rate of clipping versus endovascular repair of the aneurysm. We also defined a minimum set of parameters available: WFNS grade, diagnosis of vasospasm, clinical DCI, radiological DCI (infarct) and long-term functional outcome.

### Statistical analysis

Descriptive statistics for continuous data are presented as either mean±SD or as median and interquartile range (IQR). Categorical data were presented as counts and percentages. Comparison of continuous outcomes among two groups (e.g. Emory, SAHIT) were made using two-sample t-tests or Wilcoxon rank sum tests, as appropriate. Comparison of continuous outcomes with more than two groups was performed using ANOVA with post-hoc Scheffe testing. Nominal outcomes were compared by a Chi-square test for independence and Fisher’s exact test. Risk factors associated with binary and ordinal outcomes were assessed with multiple logistic regression or ordinal regression analysis, respectively. Results are given as odds ratios (OR) with 95% confidence intervals(CI). Propensity score-based comparative analysis,^27,28^ and the subsequent sensitivity analysis are detailed in the supplementary material.

## Results

### Patient cohort

A total of 1,351 patients were discharged from Emory University Hospital neurosciences ICU with non-traumatic SAH between 2012-2017. Of these, 993 (73.5%) were aneurysmal and 358 (26.5%) were idiopathic. Demographic information regarding the cohort is detailed in table 1. In this cohort, 36.2% (n=489) were diagnosed with clinically relevant cerebral vasospasm by means of clinical change and blood flow velocity increase on TCD, or by angiographic evidence on CTA or DSA reports. Of the patients diagnosed with vasospasm, 85.8% (n=422) were treated with IT nicardipine (Table 1). In the group that was diagnosed with vasospasm and not treated with IT nicardipine, 67.1% (n=47) did not have an EVD to allow its administration.

### Administration of IT nicardipine

Patients treated with IT nicardipine were delineated from patients who did not receive the treatment by the known risk factors for cerebral vasospasm (e.g. sex, age, smoking status, higher-grade hemorrhage, and aneurysmal etiology, Table 1). Treatment with IT nicardipine began on mean post admission day 5.8±2.8 and lasted for a total of 8.3±5.1 days. The median number of doses of IT nicardipine was 20 (IQR 18). Initial dose was 4 mg every 8 hours in 75.4% of the cases (range of 2mg every 12h to 4 mg every 4 hours). In 79.6% of the cases, the initial regimen chosen was not increased. In case of suboptimal response, or temporary response, defined by either clinical exam, or radiologic worsening on TCD or CTA, the dose was increased in 1.5% (e.g. from 2mg to 4mg), and the frequency increased in 19.5% of the cases (e.g. from q8hr to q6hr). This change, if it occurred, happened on average on day 2.6±2.3 of the treatment. Out of a concern for abrupt discontinuation promoting a rebound effect, the local practice pattern was to wean off of treatment in 75.1% of cases over 2-4 days.

### Change in proximal artery vasospasm

TCD data was available for 198 of 422 patients (46.9%) who received treatment with IT nicardipine, and 338 patients (36.4%) who did not develop clinically significant vasospasm (Figure 1A). On the day of IT nicardipine treatment initiation, the maximal mean middle cerebral artery (MCA) velocity was over 100m/s in 79.5% of the patients (CI 95%[73.2-85.8%]), above 90m/s in the anterior cerebral artery (ACA) in 74.4% [67.5%-81.2%] and above 70m/s in the basilar artery (BA) in 47.7% [39.8-55.6%]. When available, CTA-based diagnosis of vasospasm was also analyzed which demonstrated moderate to severe vasospasm in 82.8% of the treated patients (See supplementary methods and Supplementary Figure I).

**Figure 1:**
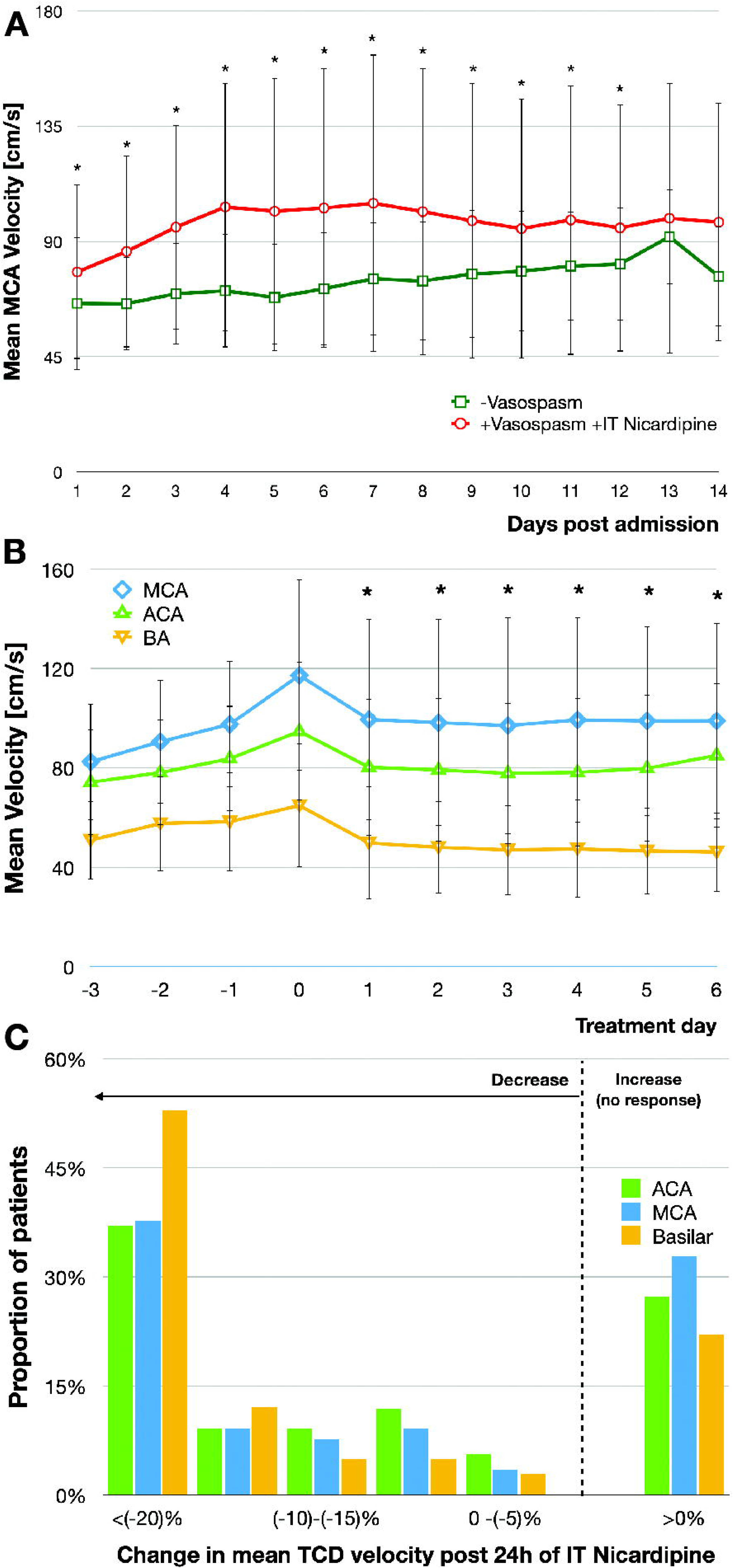
Change in blood flow velocities measured by TCD, in response to IT nicardipine. For each artery (MCA, ACA, BA), the average measurements of the mean velocity at the different depths was calculated. Results are presented as mean±SD. (A) The timeline of cerebral vasospasm: comparison of the average mean MCA velocities between the group diagnosed with vasospasm and treated with IT nicardipine, and the group that did not have clinically relevant vasospasm (* - p<0.05 between groups). (B) Change in average mean velocity in comparison to the initiation day of IT nicardipine by artery (day 0, * - p<0.05 compared to day 0). (C) Distribution of the magnitude of response in blood flow velocities within the first day of treatment (change in mean velocities between day 0 and day 1, per artery, in percentage) in patients treated with IT nicardipine. TCD – transcranial doppler; ACA – anterior cerebral artery; MCA – middle cerebral artery; BA – Basilar artery.

Treatment was associated with a reduction of mean velocities in all main arteries (ACA, MCA, BA) after one day of treatment (Relative reduction of 12.9% on average in the MCA, 13.0% in the ACA and 19.3% in the basilar territory, p<0.01, Figure 1B). This effect remained through the treatment, until the velocities normalized and plateaued. When analyzing the response in treated patients, more than 77.3% of patients showed at least one of the three main arteries experienced TCD velocity decrease within 24h of treatment (Figure 1C).

Increased TCD velocities were correlated to the risk of DCI (Figure 2). A positive response to IT nicardipine by means of decreasing blood velocities rates, however, did not correlate neither to decreased incidence of DCI nor improvement in long-term functional status (Figure 3).

**Figure 2:**
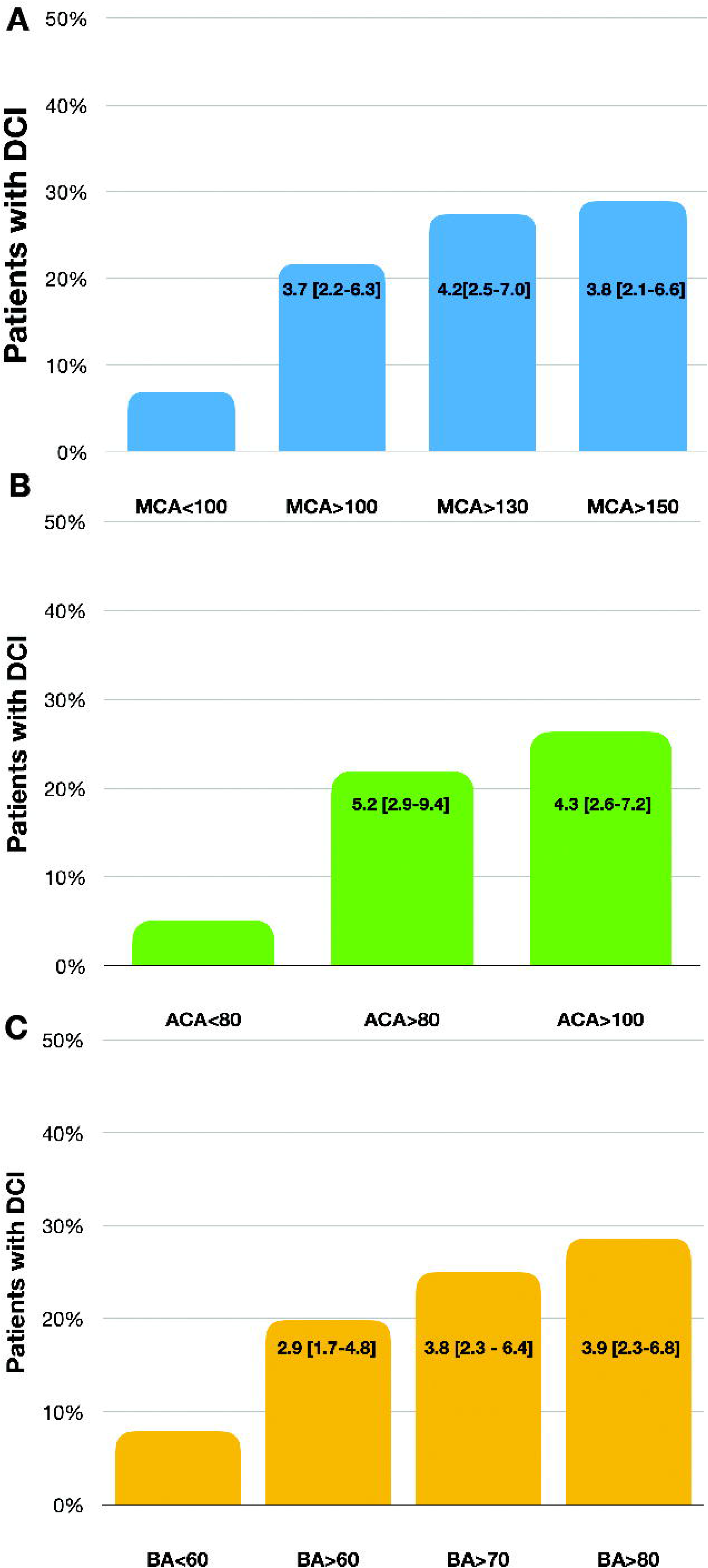
Correlation between TCD velocity and outcome. (A) Proportion of DCI according to the maximal MCA velocity (n=162). OR [CI 95%] based on unadjusted binary regression analysis is presented. (B) Proportion of DCI according to the maximal ACA velocity (n=162). (C) Proportion of DCI according to the maximal BA velocity (n=158).

**Figure 3:**
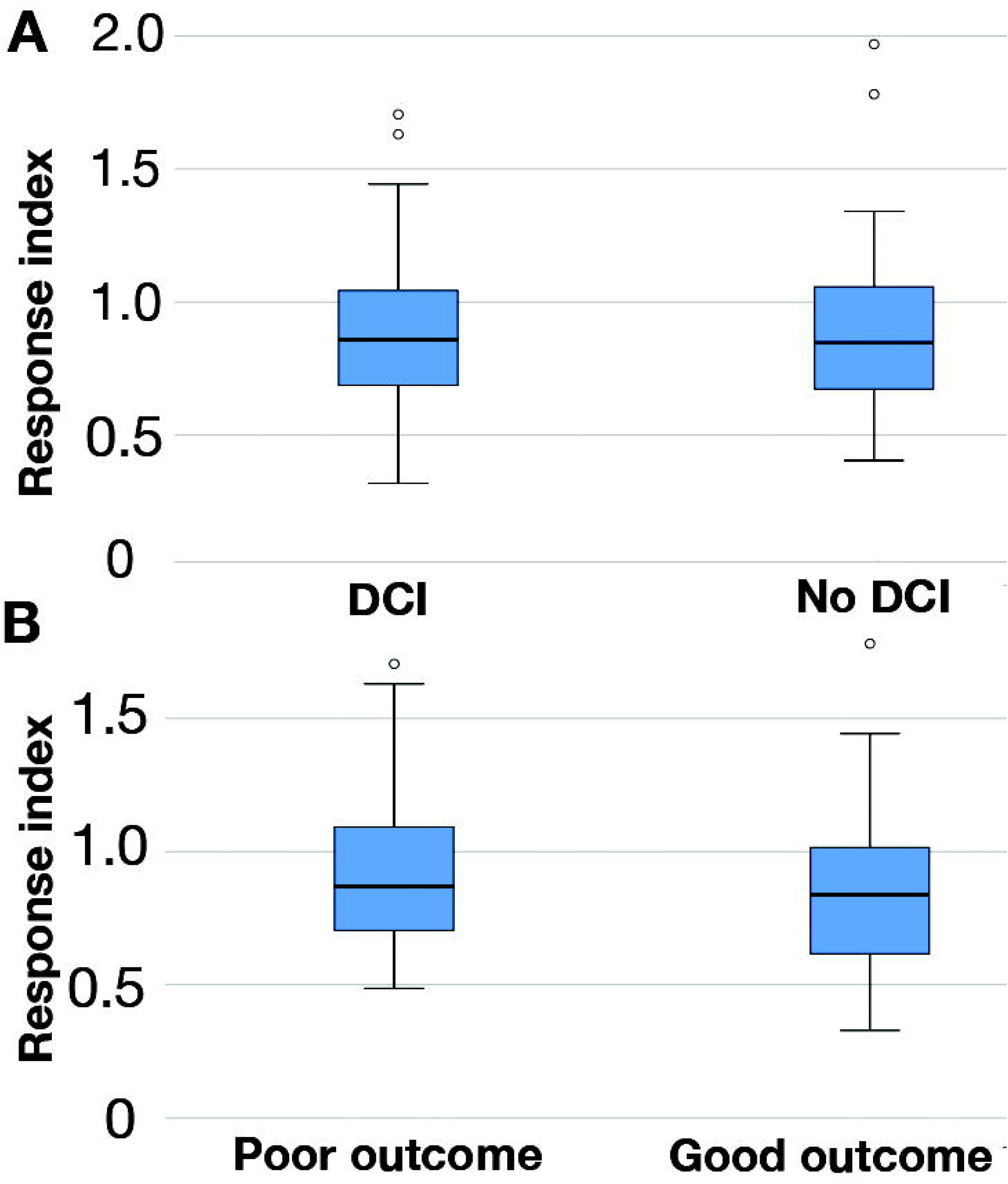
TCD MCA response index and correlation to clinical outcomes. A decrease in mean velocity in response to treatment will be noted as index<1. (A) Response index (mean MCA velocity on day one of treatment/day 0) was compared in patients with and without DCI demonstrated no statistically significant difference (n=162). (B) Response index showed no statistically significant difference in comparing patients with good long-term functional outcome (mRS≤2) with the poor outcome group (mRS>2, n=123) [Blue box represents Q1-3].

### Safety

Bacterial ventriculitis was diagnosed in 2.9% of the patients who required an EVD in our cohort (n=866). The prevalence of ventriculitis rate in patients who were treated with IT nicardipine was similar to those who did not (3.1% vs. 2.7%, respectively, OR 1.15[0.52-2.54]).

Ventriculoperitoneal shunt (VPS) was required because of persistent hydrocephalus in 14.2% in patients who had an EVD placed. Among patients who were treated with IT nicardipine, required a VPS at higher rates, compared with those who did not (19.9% vs. 8.8%, p<0.01). In a multiple logistic regression analysis, adjusting for demographics and risk factors, IT nicardipine administration was associated with an increased need for a VPS (OR 2.54 CI 95%[1.68-3.83]).

### Comparative outcome analysis

The outcome analysis of the Emory cohort is detailed in supplementary table I. Due to the retrospective nature of this study, a relevant control group within the cohort was not available. Therefore, we next performed a comparative analysis with patients from the SAHIT database. As detailed in the supplementary methods and results, following propensity score average treatment effect for treated (ATT)-based weighing, we analyzed data from 446 patients of the SAHIT dataset who were similar to the 422 patients from the Emory cohort who were treated with IT nicardipine (supplementary table I). The group treated with IT nicardipine had a lower rate of DCI, higher proportion of patients with favorable long-term functional outcome, and lower rates of bacterial ventriculitis (Table 2). The proportion of patients requiring VPS in the weighted SAHIT group was 28.4%, which was higher compared to the IT nicardipine group. Since information regarding VPS was available for only 25.1% of the SAHIT cohort, we did not include it in the logistic regression analysis.

## Discussion

This study is a retrospective description of the largest cohort to date involving the use of IT nicardipine in patients with non-traumatic SAH to treat cerebral vasospasm and prevent DCI.

The results corroborate known risk factors for cerebral vasospasm and DCI, including: younger age, women, tobacco abuse, higher WFNS and mFG. Following comparison with the SAHIT database, the data indicates that for patients diagnosed with clinically relevant vasospasm, treatment with IT nicardipine reduced proximal artery vasospasm, was associated with a lower rate of DCI and with a higher rate of good functional outcome. Moreover, IT nicardipine was not associated with higher infection rates, yet it was associated with higher rate of VPS.

Historically, vasospasm was noted on diagnostic cerebral angiography following subarachnoid hemorrhage in proximal arteries.^29^ Pathophysiology of vasospasm, still to this day, remains incompletely understood, is complex and includes vasoconstriction in part related to hemoglobin from hemolyzed red blood cells^30^, nitric oxide and endothelin pathways.^31^ For the bedside clinician identifying and intervening upon cerebral vasospasm is challenging. Radiological evidence for vasospasm is common in SAH patients. A correlation exists between vasospasm and later development of DCI. Multiple interventions in the past, however, demonstrated improvement of vasospasm without preventing DCI, nor functional outcome.^17,32–34^ In hindsight, several commonalities emerge from reviewing prior negative studies: in most studies, the treatment was prophylactic, and perhaps one dose does not fit all^35,36^; another issue is targeting resolution of vasospasm in earlier phase studies, and later failing in phase III studies when patient outcomes are the clinical end point. That was most notably relevant for the recent NEWTON II study. In NEWTON II trial, a single dose slow-release Nimodipine was administered through an EVD. The study was stopped prematurely when a planned interim analysis suggested the study had a low likelihood of meeting its primary outcome. However, it did demonstrate reduced rate of proximal vessel vasospasm, but not improvement in outcome. A trend towards improvement was noted in the more severely ill patients, suggesting that at least in part, this failure was related to patient selection.^17^ To summarize, to date, there is no proven treatment available that results in reduced vasospasm, reduce DCI and improved patient outcome in patients with cerebral vasospasm from non-traumatic SAH.^13,37^

Our experience builds on these lessons, with a treatment regimen focused on reactive, not prophylactic, treatment driven by patient response to a given treatment regimen, which is similar to prior smaller series reported.^38^ IT nicardipine in this report was largely triggered by radiological data (TCD, CTA, or DSA). Given the correlation of radiographic spasm with DCI (Figure 2), this approach likely improved patient selection for high-risk ones. The dose and length of treatment were titrated to effect, which was determined by improved neuro exam and/or improved radiographic vasospasm. This approach however introduces issues arising the spectrum of clinical decision making in a retrospective, non-protocolized cohort. As such, the definition of clinically relevant vasospasm varied between clinicians. Indeed, the majority of patients demonstrated radiological signs of mild to moderate vasospasm in more than one vessel when IT nicardipine treatment initiated. In some cases, however, initiation began with less severe spasm than others and in different distributions (one focal severe stenosis, versus milder, yet widespread vasospasm).

Cerebral vasospasm is thought to occur on both the macro and microvascular levels. Most retrospective results so far^39–41^, including several systemic reviews,^38,42,43^ suggested beneficial effect of IT nicardipine on macrovascular blood velocities, similar to our report. However, evidence has accumulated that loss of microvasculature autoregulation and consequent decrease in regional blood flow are also related to DCI.^44^ Whether IT nicardipine has an effect on the microcirculation, similar to the one it has on the proximal vessel level, is unclear. One attempt to measure the microcirculatory effect measured the effect of IT nicardipine on tissue oxygenation (PbO2) and metabolites (by microdialysis), which did not demonstrate a robust effect.^45^ The effort was limited, however, by the local measurement and probe location. The observed clinical benefit in the current report (Table 2) did not correlate with improvement in macrovascular vasospasm (Figures 1,3) suggesting that IT nicardipine may improve functional outcomes by modifying microvascular vasospasm.

We also attempted to identify complications from frequently accessing an EVD. The overall rate of bacterial ventriculitis was quite low in our cohort compared to the SAHIT one, although no routine intrathecal intervention occurred in the SAHIT cohort. Even when examining the rate of infection within our cohort, moreover, and comparing those who were treated with IT nicardipine versus those who were not, the IT treatment did not contribute to increased rate of infection. Prior reports regarding IT nicardipine treatment described an infection rate of about 6%, nearly double those reported in this publication.^43^ Whether the relatively low rate of infection is reproducible, and what is the true added risk of bacterial ventriculitis with the addition of IT nicardipine treatment remains unclear. The rate of VPS in our cohort was also lower compared with the SAHIT one, although too many data points were missing to compare the two comprehensively. In our cohort, a clear association was seen between IT nicardipine and the further need for a VPS, even after controlling for the relevant risk factors. At this time, it is unclear whether this effect is a result of an intrathecal intervention per-se, a reaction to nicardipine, or its formulation, or other explanations. Future studies that compare nicardipine with a placebo may shed light on this question.

This study has limitations arising from its retrospective nature and lack of being protocolized. When compared with our own group’s prior description of this intervention, over the course of several years, the threshold for initiation seems lower,^39^ and the overall definition of clinically significant vasospasm varied. In lieu of an internal control group we used an external database. The SAHIT dataset includes diverse cohorts, which include prior clinical trial data and single center cohorts. To reduce possible bias, we did not take into account patient cohorts that preceded the ISAT trial,^26^ since following that study, the rate of endovascular treatment for securing aneurysms became more abundant and potentially reduced rates of DCI. On the one hand, this database is based on non-protocolized practice, and therefore could have biases. On the other hand, this diversity could mean that the patient data is closer to real-life experience, similar to our cohort.

So far, prior reports and systematic reviews^38,42,43^ regarding IT Nicardipine lacked long-term outcomes. Our report suggests that a beneficial long-term effect may exist. Despite the significant steps taken to avoid bias such as using propensity-score based comparison, this analysis cannot replace a prospective controlled trial.

## Conclusion

The current study describes the largest SAH patient cohort to date that received IT nicardipine treatment to alleviate vasospasm and reduce DCI. The data shows that this treatment was associated with a reduced rate of DCI and increased proportion of patients with long-term good functional outcome. The treatment was also associated with increased rate of VPS. The results are limited by their retrospective nature. Because of the paucity of effective and proven treatments for cerebral vasospasm and prevention of DCI, these results hold promise that will need to be verified in a prospective, randomized, controlled clinical trial to test the efficacy of IT nicardipine treatment for post-SAH cerebral vasospasm.

## Data Availability

The data that support the findings of this study are available from the corresponding author upon reasonable request.

## Acknowledgments

The authors would like to thank the SAHIT collaborators (Jeroen Boogaarts [Radboud University Medical Center], Walter van den Bergh [University Medical Center Utrecht], Hitoshi Fukuda [Kurashiki Central Hospital], Daniel Hanggi [Heinrich Heine University], Peter Le Roux [University of Pennsylvania], Benjamin Lo [University of Toronto], Stephan Mayer [Columbia University], Andrew Molyneux [Oxford University], Audrey Quinn [The General Infirmary, Leeds], Gabriel Rinkel [University Medical Center Utrecht], Tom Schweizer [University of Toronto], Jose Suarez [Johns Hopkins University], Michael Todd [University of Iowa], Mervyn Vergouwen [University Medical Center Utrecht], and George Wong [Chinese University of Hong Kong) who created the comparative database used as the comparative group in this study.

## Supplemental Materials

- Supplemental methods
- Online figures:2
- Online tables:4

## Table legends

<u>Table 1:</u> Patient and clinical characteristics of the Emory Cohort. Results presented as n(%). (+ vasospasm indicates a clinical diagnosis of vasospasm; +IT nicardipine indicates the use of this intervention; WFNS – World Federation of Neurosurgical Societies; mFG – modified Fisher Grade).

<u>Table 2:</u> Clinical outcomes comparison between the IT Nicardipine treated group and the weighted group from the SAHIT dataset.

