## Supplementary material for "Intrathecal Nicardipine for Cerebral Vasospasm Post Subarachnoid Hemorrhage–a Retrospective Propensity-Based Analysis"

Assessment of severity of vasospasm by CTA

The common CTA report detailed vasospasm severity (none, mild, moderate or severe) across eleven arteries (Bilateral MCA, ACA, PCA, ICA, Vertebral and the Basilar artery). The severity was graded by neuroradiology at Emory University Hospital as part of the routine report, and the results were obtained from the official interpretation in the patients' charts. No strict numeric criteria applied to this definition, since the read was done across many years and many different neuroradiologists as a routine read and not as a study protocol.

A CTA was done in 267/422 of the nicardipine-treated patients on the day of initiation. The decision to obtain a CTA was done by the treating team (neurocritical care and neurosurgery) at real time. CTA demonstrated vasospasm at least one artery in 95.1% of the cases (Supplementary Figure IA). When taking into account only moderate to severe vasospasm, 82.8% (n=221) of the patients demonstrated it in at least one artery on day of treatment initiation. For comparison, we analyzed patients who were not diagnosed with vasospasm by the treating team and had a CTA performed at 5±2 days post admission (day 5 is the median day of IT nicardipine initiation, n=149). In this group 72.5% did not have moderate-severe vasospasm noted in any artery.

Since vasospasm was often multifocal, and severity varied significantly, we attempted to score the overall severity of the angiographic vasospasm. The written interpretation across 11 vessels was scored: vasospasm for each artery was graded as 0 for no spasm; 1 for mild; 2 for moderate; and 3 for severe. By summing across 11 vessels, each CTA could be scored within the range of 0-33 points in this scale. We summarized the score of CTAs with patients who had a CTA done on the day of IT nicardipine initiation (n=267) and compared it with patients who were not treated for vasospasm and had a CTA performed on day 5±2 (n=149).

Comparison of the two groups demonstrated a median score of 9 (Q1-Q3[5-14]) in the IT nicardipine group and 1[0-4] in the group that was not treated with IT nicardipine (p<0.01). 73.15% of the patients who were assessed not to have a clinically relevant vasospasm, had a very low score of 3 or below, compared with 17.23% in the group that was treated with IT nicardipine (Figure IB-C).

**Figure I**: Vasospasm score by CTA. (A) Histogram detailing the proportion of patients diagnosed with moderate to severe vasospasm by the radiology CTA report, by number of arteries involved. The comparison done between patients who were diagnosed with vasospasm and treated with IT nicardipine (+IT nicardipine group) versus the group who did not develop clinically significant vasospasm, yet a CTA was available from hospital day 5±2 (median day of IT nicardipine initiation). (B) CTA-Vasospasm score for patients who were not diagnosed with clinically relevant vasospasm and had a CTA done on day 5±2 of admission. (C) CTA-Vasospasm score for patients who were treated with IT nicardipine from the day of treatment initiation.

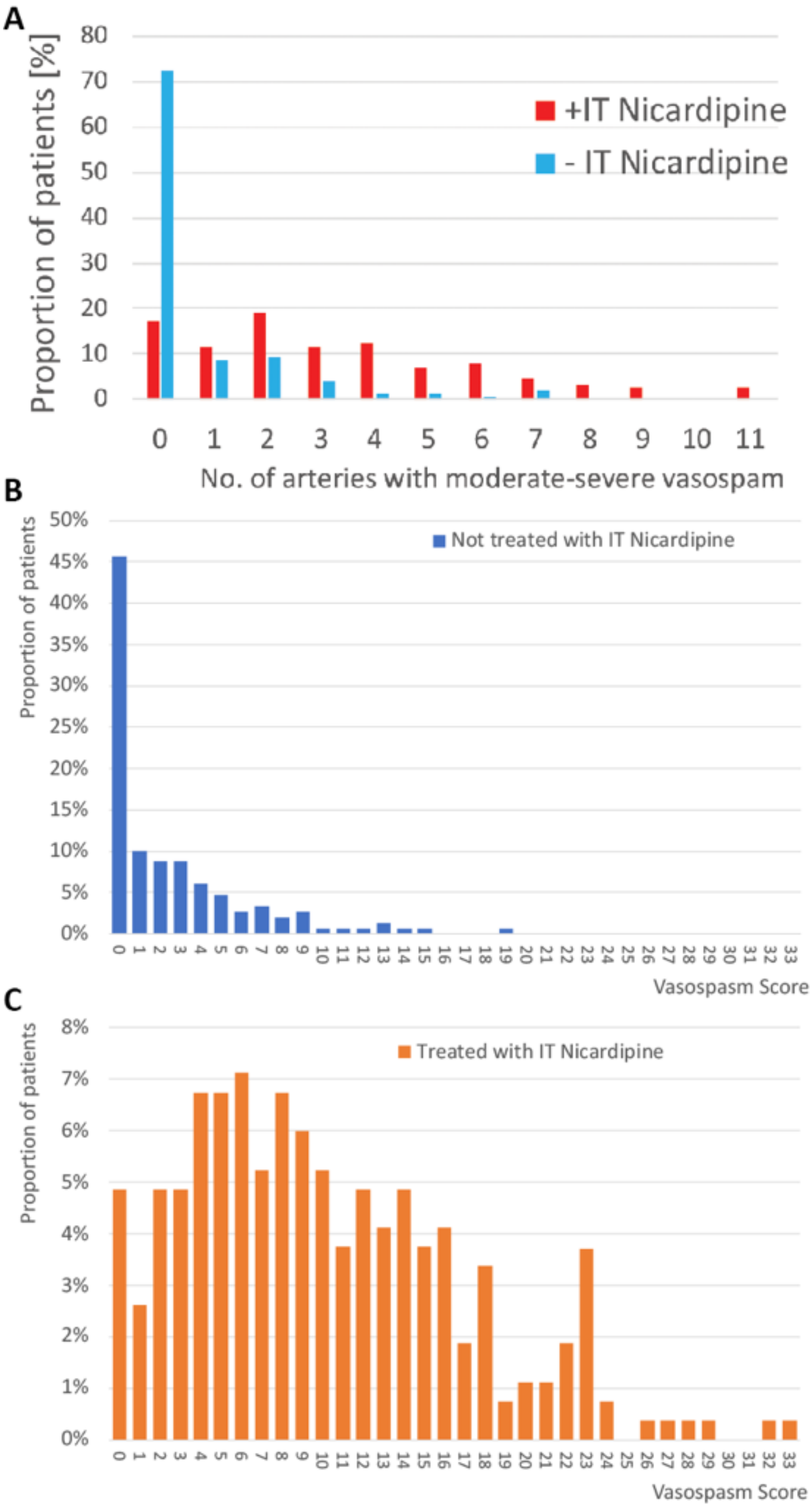

A
Proportion of patients [%]
+IT Nicardipine
- IT Nicardipine
No. of arteries with moderate-severe vasospam
B
Proportion of patients
Not treated with IT Nicardipine
Vasospasm Score
C
Proportion of patients
Treated with IT Nicardipine
Vasospasm Score

Emory Cohort Patient Outcomes

| | ***WFNS group*** | ***All*** | ***-Vasospasm - IT nicardipine*** | ***+Vasospasm -IT nicardipine*** | ***+Vasospasm +IT nicardipine*** |
|---|---|---|---|---|---|
| *N* | | 1351 | 859 | 70 | 422 |
| *DCI [%(N)]* | WFNS 1 | 4.8% (601) | 2.2% (448) | 2.0% (49) | 17.3% (104) |
| | WFNS 2 | 11.4% (272) | 5.5% (146) | 0% (9) | 19.8% (116) |
| | WFNS 3 | 9.7% (72) | 7.1% (42) | 50% (2) | 10.7% (28) |
| | WFNS 4 | 17.4% (224) | 12.4% (97) | 0% (6) | 22.3% (121) |
| | WFNS 5 | 10.6% (180) | 3.3% (123) | 25% (4) | 26.9% (52) |
| | All | 9.3% (1,347) | 4.3% (856) | 4.3% (70) | 20.2% (421) |
| *In patient mortality [%(N)]* | WFNS 1 | 3.0% (603) | 2.3% (450) | 0% (49) | 7.7% (104) |
| | WFNS 2 | 7.3% (272) | 8.3% (146) | 0% (9) | 6.8% (117) |
| | WFNS 3 | 11.1% (72) | 14.3% (42) | 0% (2) | 7.2% (28) |
| | WFNS 4 | 22.8% (224) | 35.1% (97) | 0% (6) | 14.0% (121) |
| | WFNS 5 | 74.5% (180) | 92.7% (124) | 50% (4) | 32.7% (52) |
| | All | 17.1% (1,351) | 20.2% (859) | 2.9% (70) | 12.3% (422) |
| *Long-term functional status by mRS (Median, IQR, N)* | WFNS 1 | 1 (0-1, 477) | 1 (0-1, 347) | 0 (0-1, 40) | 1 (1-2, 90) |
| | WFNS 2 | 1 (0-2, 219) | 1 (0-2, 119) | 1 (1-2, 8) | 1 (1-3, 92) |
| | WFNS 3 | 2 (1-3, 58) | 2 (1-4, 32) | 3(3-3,1) | 1 (1-2, 25) |
| | WFNS 4 | 3 (1-6, 166) | 4 (1-6, 80) | 2 (1-3,5) | 3 (1-5, 81) |
| | WFNS 5 | 6 (6-6, 163) | 6 (6-6, 122) | 6(2-6,4) | 5 (3-6, 37) |
| | All | 1 (1-3, 1,083) | 1 (0-6, 700) | 1(0-1, 58) | 2(1-4,325) |

**Table I**: Outcomes of the EUH patient cohort by groups and WFNS. The -Vasospasm, - IT nicardipine group was not clinically diagnosed with cerebral vasospasm; the +Vasospasm, -IT nicardipine group was clinically diagnosed with vasospasm, but was not treated with IT nicardipine. The +Vasospasm +IT nicardipine was both diagnosed with vasospasm and treated with IT nicardipine. Results are calculated for existing data, excluding missing ones. For all analysis the N used is mentioned. Of note, the group treated with IT nicardipine was the group with highest risk for DCI, and indeed had the highest proportion of patients with DCI, and respectively worse functional outcome. These results, however, are without an adequate comparison group, and therefore the propensity score analysis comparing this cohort to an external one, occurred (see below and in the main text).
The group that had no clinically relevant vasospasm comprises two major populations: patients who indeed had no vasospasm, and a relatively benign course, and a group that was very sick and did not survive long enough to develop vasospasm. Indeed, the mortality rate in the WFNS 4-5 subgroup without vasospasm was higher compared to those who had vasospasm and treated with IT nicardipine.

Propensity score detailed methods

Comparative analysis between our cohort and the SAHIT took place after dichotomizing the dataset into two groups (1) Emory patients with diagnosed cerebral vasospasm and were treated with IT nicardipine (+vasospasm +IT nicardipine group); (2) SAHIT patients with diagnosed cerebral vasospasm not treated with IT nicardipine. To our knowledge, no patients in the SAHIT database were treated with IT nicardipine. To avoid bias, we specifically excluded the intervention group from the NEWTON-2 trial because they were treated with slow-release preventive IT Nimodipine. Given the non-randomized design of the current study, patients who were treated IT nicardipine for cerebral vasospasm might have differed demographically and clinically from those that do not. Differences in underlying demographic and clinical characteristics (e.g., age, sex, race/ethnicity, WFNS score, cardiovascular risk factors, neurosurgical approach to the aneurysm repair etc.) were identified by comparing the two cohorts using two-sample t-tests, Wilcoxon rank sum tests, or Chi-square tests, as appropriate. After analyzing the completeness of the data, we removed race as a covariate (29.4% present) and hypercholesterolemia/dyslipidemia (30.7% present). We included the following variables in the propensity score (PS) model, which were common to the EUH and SAHIT datasets: age, gender, diabetes mellitus, smoking status, hypertension, coronary artery disease, surgical treatment, WFNS scale, and modified Fisher Grade. Before weighting, all variables were found to be statistically different except for diabetes mellitus and coronary artery disease (Table II). Variables identified as significantly different in this bivariate analysis were used to adjust the association between IT nicardipine use and clinical outcomes of interest (DCI present/absent and favorable outcome at 3mo, defined as mRS≤2 or GOSE>6). To allow for the adjustment of baseline differences between the two cohorts and to reduce the possible effects of confounding variables, we utilized propensity score methods to balance treatment cohorts on demographic and clinical characteristics that may affect outcome measures. First, using a logistic regression model, we regressed the treatment (IT nicardipine use (yes vs. no)) on age, gender, diabetes mellitus, smoking status, hypertension, coronary artery disease, surgical treatment, WFNS scale, and modified Fisher Grade. Probabilities associated with the likelihood of receiving IT nicardipine, conditional on baseline characteristics, were calculated for each patient. Probabilities were converted to average treatment effect on treated weights (ATT). Once the ATT weights were finalized, at the second step, adjusted analyses examined the effect of IT nicardipine treatment on each outcome of interest (DCI, outcome at 3mo) using the =ATT weight in all statistical models. For each outcome of interest, we fit a multivariable logistic regression model using double robust propensity score methods. Each patient was weighted by their ATT weight, and the treatment group and logit of the propensity score were included as the covariates in this second model.

To estimate the propensity score, a logistic regression model with IT nicardipine as the outcome variable was fit with predictor variables age, gender, diabetes mellitus, smoking status, hypertension, coronary artery disease, hypercholesterolemia/dyslipidemia, surgical treatment, WFNS scale, and modified Fisher Grade. The initial propensity scores are plotted for each dataset in figure IIA. Improvement in propensity score overlap after ATT weighting is shown in figure IIB, and improvement in the distribution of baseline covariates by group is seen in table III (weighted). The primary criteria for evaluating our propensity score weighting was met, namely that the baseline covariates are balanced between groups.

**Table II. Baseline Characteristics of all Emory patients and the SAHIT patients**

| | *All EUH* | *SAHIT* | *P-Value for Difference* |
|---|---|---|---|
| *N* | 1351 (100.00) | 4986 | |
| *Age* | 54.46 (13.82) | 55.77 (13.78) | 0.002 * |
| | 55 (46, 63) | 55 (47,65) | 0.007 * |
| *Gender* | | | 0.013 * |
| ***Female*** | 906 (67.06) | 3416 (70.62) | |
| ***Male*** | 445 (32.94) | 1421 (29.38) | |
| *Race/ethnicity* | | | <0.001 * |
| ***African American*** | 452 (33.46) | 19 (9.89) | |
| ***White*** | 581 (43.01) | 148 (77.08) | |
| ***Asian*** | 34 (2.52) | 16 (8.33) | |
| ***Hispanic/Latino*** | 39 (2.89) | 0 (0.00) | |
| ***Other/Unknown*** | 224 (16.58) | 9 (4.69) | |
| *Diabetes* | | | <0.001 * |
| ***Yes*** | 167 (12.36) | 180 (7.24) | |
| ***No*** | 1184 (87.64) | 2307 (92.76) | |
| *Smoker* | | | <0.001 * |
| ***Yes*** | 378 (27.98) | 1347 (51.93) | |
| ***No*** | 973 (72.02) | 1247 (48.07) | |
| *Hypertension* | | | <0.001 * |
| ***Yes*** | 759 (56.18) | 1286 (45.43) | |
| ***No*** | 592 (43.82) | 1545 (54.57) | |
| *Coronary Artery Disease* | | | 0.001 * |
| ***Yes*** | 93 (6.88) | 110 (4.44) | |
| ***No*** | 1258 (93.12) | 2369 (95.56) | |
| *Hypercholesterolemia/ Dyslipidemia* | | | 0.407 |
| ***Yes*** | 216 (15.99) | 296 (17.11) | |
| ***No*** | 1135 (84.01) | 1434 (82.89) | |
| *Vasospasm* | | | 0.281 |
| ***Yes*** | 492 (36.42) | 1413 (38.08) | |
| ***No*** | 859 (63.58) | 2298 (61.92) | |
| *Surgical Treatment* | | | <0.001 * |
| ***Clip*** | 241 (17.84) | 1784 (38.99) | |
| ***Endovascular*** | 622 (46.04) | 1629 (35.61) | |
| ***Idiopathic (Angio negative)*** | 358 (26.50) | 310 (6.78) | |

| | | | |
|---|---|---|---|
| ***No Treatment*** | 130 (9.62) | 852 (18.62) | |
| *WFNS Scale* | | | 0.001 |
| ***1*** | 604 (44.71) | 1884 (39.85) | |
| ***2-3*** | 343 (25.39) | 1208 (25.55) | |
| ***4-5*** | 404 (29.90) | 1636 (34.60) | |
| *mFG* | | | <0.001 * |
| ***0*** | 26 (1.99) | 97 (2.27) | |
| ***1*** | 161 (12.32) | 580 (13.59) | |
| ***2*** | 117 (8.95) | 473 (11.08) | |
| ***3*** | 231 (17.67) | 1617 (37.90) | |
| ***4*** | 772 (59.07) | 1500 (35.15) | |

**Table III** Demographic and Summary data for all patients and patients by group, pre- and post-Average Treatment Effect on Treated (ATT) weighting. Results presented as n(%).

| | Overall | | SAHIT | | IT nicardipine | P Value | |
|---|---|---|---|---|---|---|---|
| | **Unweighted** | **Weighted** | **Unweighted** | **Weighted** | | **Unweighted** | **Weighted** |
| N | 1835 | 868 | 1413 | 446 | 422 | NA | NA |
| Age, mean(SD) | 52.8±12.4 | 51.0±9.6 | 53.3±12.4 | 51.0±8.4 | 51.1±12.4 | 0.002 | 0.77 |
| Gender | | | | | | <.0001 | 0.18 |
| **Female** | 1174 (64.0) | 651 (75.0) | 866 (61.3) | 343 (76.6) | 308 (73.0) | | |
| **Male** | 661 (36.0) | 217 (25.0) | 547 (38.7) | 103 (23.4) | 114 (27.0) | | |
| Diabetes | | | | | | 0.72 | 0.48 |
| **Yes** | 101 (7.0) | 58 (6.7) | 70 (6.8) | 27 (6.2) | 31 (7.4) | | |
| **No** | 1349 (93.0) | 809 (93.3) | 958 (93.2) | 418 (93.8) | 391 (92.6) | | |
| Smoker | | | | | | <.0001 | 0.62 |
| **Yes** | 813 (49.2) | 276 (31.8) | 674 (54.7) | 137 (30.6) | 139 (32.9) | | |
| **No** | 841 (50.8) | 592 (68.3) | 558 (45.3) | 309 (69.4) | 283 (67.1) | | |
| Hypertension | | | | | | 0.002 | 0.13 |
| **Yes** | 721 (49.5) | 504 (58.0) | 486 (46.9) | 268 (60.2) | 235 (55.7) | | |
| **No** | 737 (50.5) | 364 (42.0) | 550 (53.1) | 178 (39.8) | 187 (44.3) | | |
| Coronary Artery Disease | | | | | | 0.16 | 0.79 |
| **Yes** | 59 (4.1) | 44 (5.0) | 37 (3.6) | 22 (4.8) | 22 (5.2) | | |
| **No** | 1390 (95.9) | 824 (95.0) | 990 (96.4) | 424 (95.2) | 400 (94.8) | | |
| Surgical Treatment | | | | | | <.0001 | 0.75 |
| **Clip** | 581 (31.9) | 257 (29.6) | 453 (32.3) | 129 (29.0) | 128 (30.3) | | |
| **Endovascular** | 598 (32.8) | 544 (62.7) | 340 (24.3) | 286 (64.1) | 258 (61.1) | | |
| **Idiopathic (Angio negative)** | 128 (7.0) | 49 (5.7) | 101 (7.2) | 22 (5.0) | 27 (6.4) | | |
| **No Treatment** | 517 (28.3) | 18 (2.0) | 508 (36.2) | 9 (2.0) | 9 (2.1) | | |

| | | | | | | | |
|---|---|---|---|---|---|---|---|
| WFNS Scale | | | | | | | |
| **1** | 636 (35.3) | 212 (24.5) | 532 (38.6) | 109 (24.4) | 104 (24.6) | <.0001 | 0.35 |
| **2-3** | 504 (28.0) | 318 (36.6) | 359 (26.0) | 173 (38.7) | 145 (34.4) | | |
| **4-5** | 662 (36.7) | 337 (38.9) | 489 (35.4) | 164 (36.9) | 173 (41.0) | | |
| mFG | | | | | | <.0001 | 0.57 |
| **0-1** | 256 (14.1) | 24 (2.7) | 244 (17.5) | 12 (2.6) | 12 (2.9) | | |
| **2** | 136 (7.5) | 54 (6.2) | 107 (7.7) | 25 (5.7) | 29 (6.9) | | |
| **3** | 529 (29.2) | 107 (12.3) | 472 (33.9) | 50 (11.1) | 57 (13.5) | | |
| **4** | 892 (49.2) | 682 (78.7) | 569 (40.9) | 359 (80.6) | 323 (76.7) | | |

**Figure II**: Histogram of propensity scores by group. (A) Histogram of the propensity score of the Emory (EUH) dataset compared with the SAHIT prior to ATT weighting. (B) Post weighting, the histograms of both groups were better overlapping.

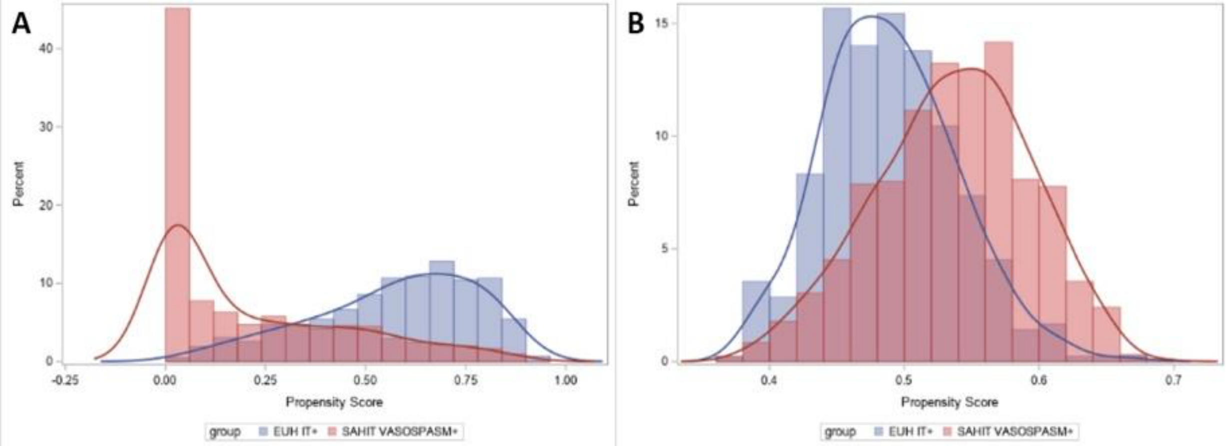


**Sensitivity analysis**

We used Average Treatment Effect on Treated (ATT) weights in the primary propensity score (PS) analysis[1]. The double robust logistic regression model included logit(propensity score) and used average treatment effect for treated weights. Propensity score model included age, gender, diabetes, smoking status, hypertension, coronary artery disease, dyslipidemia, surgical treatment, WFNS scale, and Modified Fisher Grade. Sample included 422 EUH IT nicardipine patients and 446 SAHIT patients.

To conduct our first sensitivity analysis for our results, we fit a logistic regression model for each outcome of interest without using propensity score weighting. If these results were substantially different from our conclusions using propensity score weighting, that would indicate a strong confounding effect of the observed covariates on our outcomes. The logistic regression model included treatment (EUH IT nicardipine patients versus SAHIT patients with vasospasm), age, gender, diabetes, smoking status, hypertension, coronary artery disease, surgical treatment, WFNS scale, and Modified Fisher Grade. Sample included 422 EUH IT nicardipine patients and 1413 SAHIT patients.
We used ATT weighting rather than propensity score matching in our primary analysis, as it would allow us to keep a larger sample of patients. Matching may also select a subset of

patients that are not representative of the overall patient population. Some evidence exists, however, that ATT weighting is sensitive to extreme outliers in propensity score.[44] For a second sensitivity analysis, we re-analyzed the data using propensity score matching, to see how sensitive our ATT weighted estimates were to any outliers. The regression model included group (EUH IT nicardipine patients versus SAHIT patients with vasospasm). The propensity score model included age, gender, diabetes, smoking status, hypertension, coronary artery disease, surgical treatment, WFNS scale, and Modified Fisher Grade. Sample included 421 EUH IT nicardipine patients and 421 SAHIT patients (Table IV). We concluded, based on the sensitivity analysis, that the results of our primary analysis, and that a double robust ATT-weighted propensity score model was the most appropriate method for the primary analysis.

**Table IV:** Sensitivity analysis - Odds Ratios for Intrathecal nicardipine Associated with Favorable Outcome at 3m and DCI using three different methods.

| | Odds Ratio (95% CI) | | |
|---|---|---|---|
| **Outcome** | **Double Robust ATT PS Weighted Model** | **Logistic Regression, no PS** | **Matching with PS** |
| **Favorable Outcome at 3m** | 2.17 (1.61, 2.91) | 3.30 (2.36, 4.63) | 2.39 (1.79, 3.19) |
| **DCI** | 0.61 (0.44, 0.84) | 0.55 (0.39, 0.78) | 0.59 (0.43, 0.81) |
